# Prognostic Significance of WT1 Expression Level Thershold in Acute Myeloid Leukemia Patients Receiving Hematopoietic Stem Cell Transplantation: Meta-Analysis

**DOI:** 10.1101/2022.04.18.22273981

**Authors:** Dandan Wang, Ling Qin, Boya Li, Tong Li

## Abstract

**Objective:** The WT1 gene is considered as a poor prognostic factor for acute myeloid leukemia (AML) after Allogeneic hematopoietic stem cell transplantation (Allo-HSCT). However, the effect of the expression threshold of WT1 on the prognosisis controversial, which is evaluated in this meta-analysis.

**Methods:** Relevant studies about the expression threshold of WT1 on the prognosis of AML after Allo-HSCT were searched in online databases. Data were extracted from them and analyzed by Stata16.0 software.

**Results:** Five studies involving 739 patients were screened out, including 433 cases experimental group and 306 cases control group. The experimental group and control group were compared for 1-year disease-free survival rate (DFS) [RR=1.19, 95%CI (1.03, 1.38), *P* =0.02] and 4-year DFS [RR= 1.18, 95%CI (0.98, 1.42), *P* =0.09]. The experimental group was lower than the control group in 1-year DFS, and there was no statistical significance in 4-year DFS. 1-year overall survival rate (OS) [RR=1.06, 95%CI (0.92, 1.23), *P* =0.40] and 4-year OS [RR= 1.16, 95%CI (1.03, 1.32), *P* =0.02], suggesting that the experimental group had a lower 4-year OS than the control group, and 1-year OS had no statistical significance.

**Conclusions:** High WT1 expression is unfavorable to the prognosis of AML patients undergoing Allo-HSCT. A threshold of 250 copies/104ABL of WT1 may be the best value for predicting the poor prognosis in these patients.

## Intorduction

Acute myeloid leukemia (AML), a group of heterogeneous diseases with different clinical outcomes, involves with cytogenetic and genetic lesions of hematopoietic stem cells or progenitor cells [1], increasing incidence with age[3] as the most common acute leukemia [2], for which the most effective treatment is Allogeneic Hematopoietic Stem Cell Transplantation (Allo-HSCT) [4]. However, the recurrence of patients with myeloid malignancies such as AML often causes the failure of hematopoietic stem cell transplantation, therefore, it is required to perform early monitoring of minimal residual leukemia (MRD) after Allo-HSCT[5].

First, Wilms tumor gene (WT1) involved with AML is expressed in all blast cells of acute leukemia patients, as WT1 nucleoprotein is detected in most of these blast cells [6]. More and more evidence shows that due to high expression of WT1 in various types of AML, WT1 can be used as a useful marker for monitoring MRD [7]. In addition, WT1 has a low expression in normal bone marrow, a high expression at the diagnosis of AML patients and after effective treatment, even a higher expression again before clinical recurrence [8], with higher sensitivity and specificity [5]. Although most studies have acknowledged the predictive value of WT1, few prognostic interventions have been based on WT1 expression levels [7]. Therefore, this study aims to comprehensively evaluate the influence of WT1 on the clinical prognosis of patients after Allo-HSCT by Meta-analysis method for providing a reference for clinical medical work.

## Materials and methods

Research type: Randomized controlled trials (RCT) or cohort study. Research subject: (1) AML patients receiving Allo-HSCT after chemotherapy induction and immune intervention; (2) No limitation of the age. Detection of WT1 expression: (1) WT1 expression was detected in bone marrow samples early after transplantation; (2) According to the European Leukemia Network (ELN), real-time quantitative PCR (RT-qPCR) was used to determine the mRNA levels of WT1 and ABL, and the latter (104 copies of ABL) was used as an internal reference to normalize the expression level of WT1 Interventions: According to the expression level of WT1, it was divided into high expression and low expression. Categorized by the threshold of WT1, AML patients after HSCT were further divided into experimental group (250 copies /104ABL of WT1 as the threshold) and control group (100 copies/104ABL of WT1 as the threshold). Outcomes: 1-year, 2-year, 4-year and 7-year recurrence rate (RR) and overall survival rate (OS) in high expression and low expression, and 1-year and 4-year disease-free survival rate (DFS) and overall survival rate (OS) in experimental group and the control group. Exclusion criteria: (1) Literatures with incomplete data or indicators that cannot be extracted; (2) Repeated publications; (3) Literatures not published in Chinese or English language; (4) Review, conference, abstract, case report, etc.

### Search strategy

The study based on PubMed, The Cochrane Library, Embase, CNKI, Wanfang and China Biomedical Literature Database (CBM) aims to searched for the effect of WT1 on the prognosis of AML after HSCT, with the retrieval time from the establishment of the database to December 2020, as well as references included in the study are traced back to supplement relevant literature. The retrieval of subject words, carried out by the combination with free words, is adjusted according to the characteristics of each database. In English, “WT1” and “hematopoietic transplantation” are used as search terms, while “WT1” and “hematopoietic stem cell transplantation” in Chinese.

### Literature screening and data extraction

Two researchers were independently responsible for literature screening, data extraction and cross-check. Any disagreement was solved by the third research through discussion or consultation. Briefly, literatures were initially screened by reviewing the title, followed by the abstract and full-text determined based on inclusion and exclusion criteria. Missing data were obtained by contacting the first author through e-email or telephone if necessary. The following data were mainly extracted: (1) Basic information, including the first author, country, publication date, etc. (2) Basic characteristics of subjects, including sample size, age, 250 copy threshold, 100 copy threshold, etc. (3) Outcomes: 1-year, 2-year, 4-year and 7-year RR and OS in high expression and low expression, and 1-year and 4-year DFS and OS in experimental group and the control group. If the survival curve was obtainable in literature, the survival index and recurrence index corresponding to each point were yielded using Engauge Digitizer 10.8 software. (4) Key elements of bias risks.

### Risk assessment on bias in included studies

Two researchers independently evaluate and cross-check the risk of bias in the included studies. In terms of differences, they discuss and resolve, and refer to a third party for adjudication if necessary. The RCT bias risk assessment is performed using the RCT bias risk assessment tool recommended in the Cochrane Manual 5.1.0., while the risk of bias in the included cohort studies is evaluated by the Newcastle-Ottawa Scale (NOS). The specific contents of the assessment included: 1.Selection,1)Represent activeness of the exposed cohort;2) Selection of non-exposed cohort;3)Ascertainment of exposure;4) Demonstration that outcome of interest was not present at the beginning of study.2.Comparability:1)Comparability of cohort on the basis of the design or analysis.3.Outcome:1) Assessment on outcome;2)follow-up long enough for outcomes to occur;3) Adequacy of follow-up of cohorts. The full mark of comparability between exposure and non-exposure is 2, and the remaining sub-items are all scored 1. The NOS scale scores on a scale of 9, 0-4 stands for low quality studies,5-6 for medium quality research and 7-9 for high quality research.

### Statistical analysis

Stata 16.0 software is used for Meta analysis. Risk ratio (RR) with 95% confidence interval (95%CI) is used as effect size for classification variables. The heterogeneity among the included results is analyzed by χ^2^ test (the test level was α=0.1), and the magnitude of heterogeneity is quantitatively determined by I^2^. P<0.10 or I^2^≥50% indicates significant heterogeneity. If there is no statistical heterogeneity among the results, the fixed-effect model will be used for Meta-analysis. If there is statistical heterogeneity among the study results, the random effects model will be used for Meta-analysis, and the sensitivity analysis conducted by excluding references one by one. All publication offsets are first judged by funnel plot, and then Egger test is used to evaluate the possibility of offsets. P<0.05 indicates that the difference is statistically significant. TSA 0.9 software was used for sequential analysis to evaluate the sample size of meta-analysis and verify the accuracy of the results.

## Results

### Literature screening and meta-analysis results

A total of 1460 literatures were initially screened out. After thoroughly reviewing abstracts and full-text, a total of 739 patients in 5 cohort studies were involved in the present study [9-13], including 217 patients with high expression and 522 patients with low expression. Classified by the expression threshold of WT1, there were 433 cases in the experimental group with the threshold of 250 copies/104ABL, and 306 cases in the control group with the threshold of 100 copies/104ABL. The literature screening process and their information were shown in Figure1.

**Figure 1:**
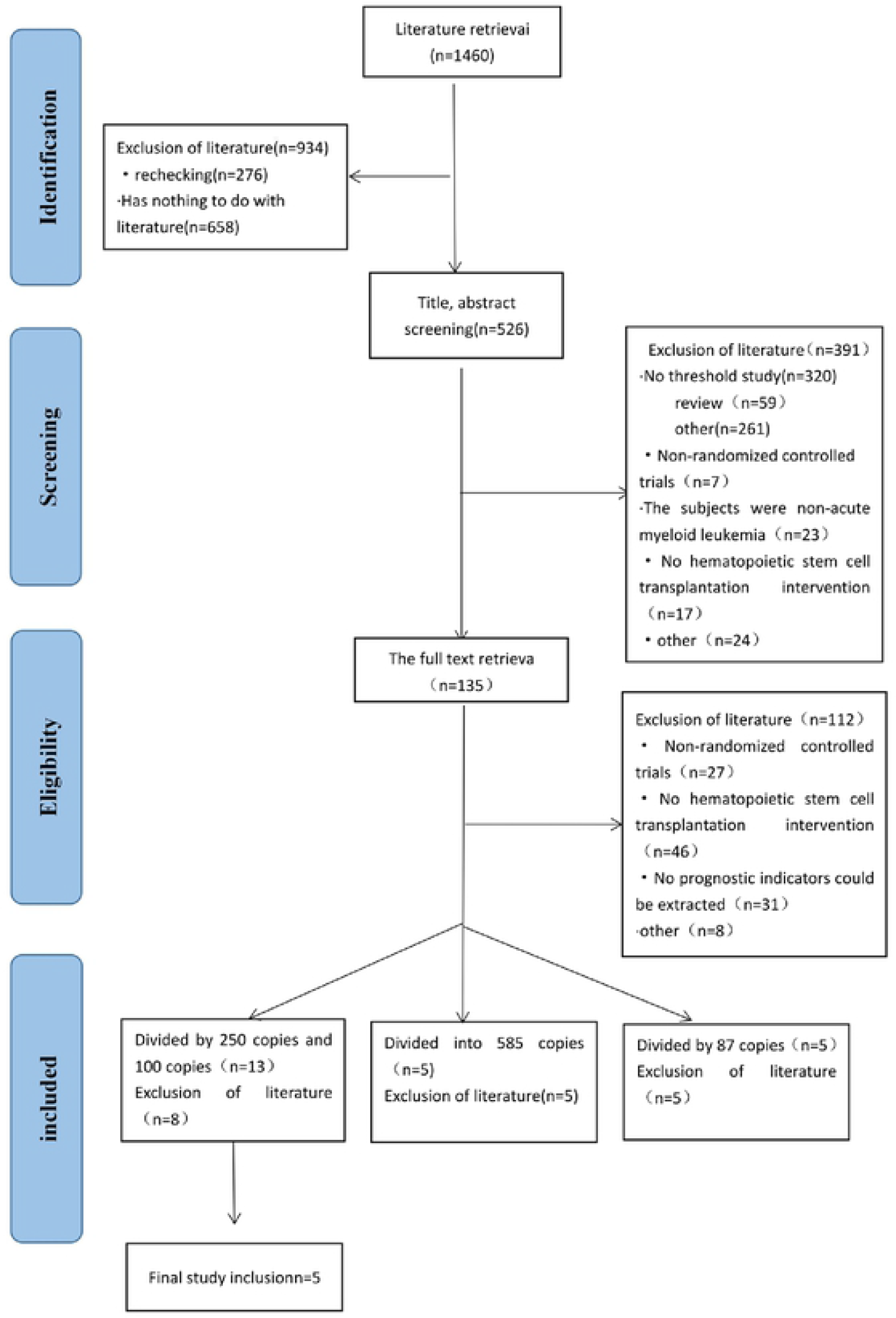
The Preferred Reporting Items for Meta-Analyses flow chart for study selection.

### Basic characteristics of the included studies and quality evaluation of included studies

The basic characteristics of included studies and WT1 expression are shown in Table 1and Table 2. Those 5 included studies are all cohort studies. Literature quality score shows that high quality of the included studies, as shown in Table 3.

**Table 1:** Characteristics of the included studies.

| The first author | Year | Country | WT1 expression<br>threshold (/$10^4$<br>ABL) | Sample<br>size | Age | Outcome indicat<br>or | Quality Score |
| --- | --- | --- | --- | --- | --- | --- | --- |
| Josep F. Nomde[9] | 2018 | Spain | 100copies | 184 | All | DFS、 OS | 8 |
| Sarah Pozzi[10] | 2013 | Italy | 100copies | 122 | 15-69 | OS | 9 |
| Anna Candoni[11] | 2017 | Italy | 250copies | 122 | 18-70 | DFS、 OS | 8 |
| Anna Candoni[12] | 2017 | Italy | 250copies | 54 | 20-69 | DFS、 OS | 8 |
| Silvia Park [13] | 2020 | Korea | 250copies | 257 | 15-75 | — | 9 |

Abbreviations: AML,Acute Myelogenous Leukemia; RR, Recurrence Rate; OS,Overall Survival; DFS,Disease Free Survival.

**Table 2:** Characteristics of WT1 expression.

| Included studies | Experimental group (250 copies /104 ABL) |  |  | Control group (100 copies /104 ABL) |  |  |
| --- | --- | --- | --- | --- | --- | --- |
|  | high expression | low expression | outcome indicator | high expression | low expression | outcome indicator |
| Josep F. Nomde2018[9] | — | — | — | 32 | 152 | — |
| Sarah Pozzi2013[10] | — | — | — | 67 | 35 | RR、 OS |
| Anna Candoni2017[11] | 41 | 81 | RR、 OS | — | — | — |
| Anna Candoni2017[12] | 21 | 33 | RR、 OS | — | — | — |
| Silvia Park2020 [13] | 56 | 201 | RR、 OS | — | — | — |

Abbreviations:RR, Recurrence Rate; OS,Overall Survival.

**Table 3:**
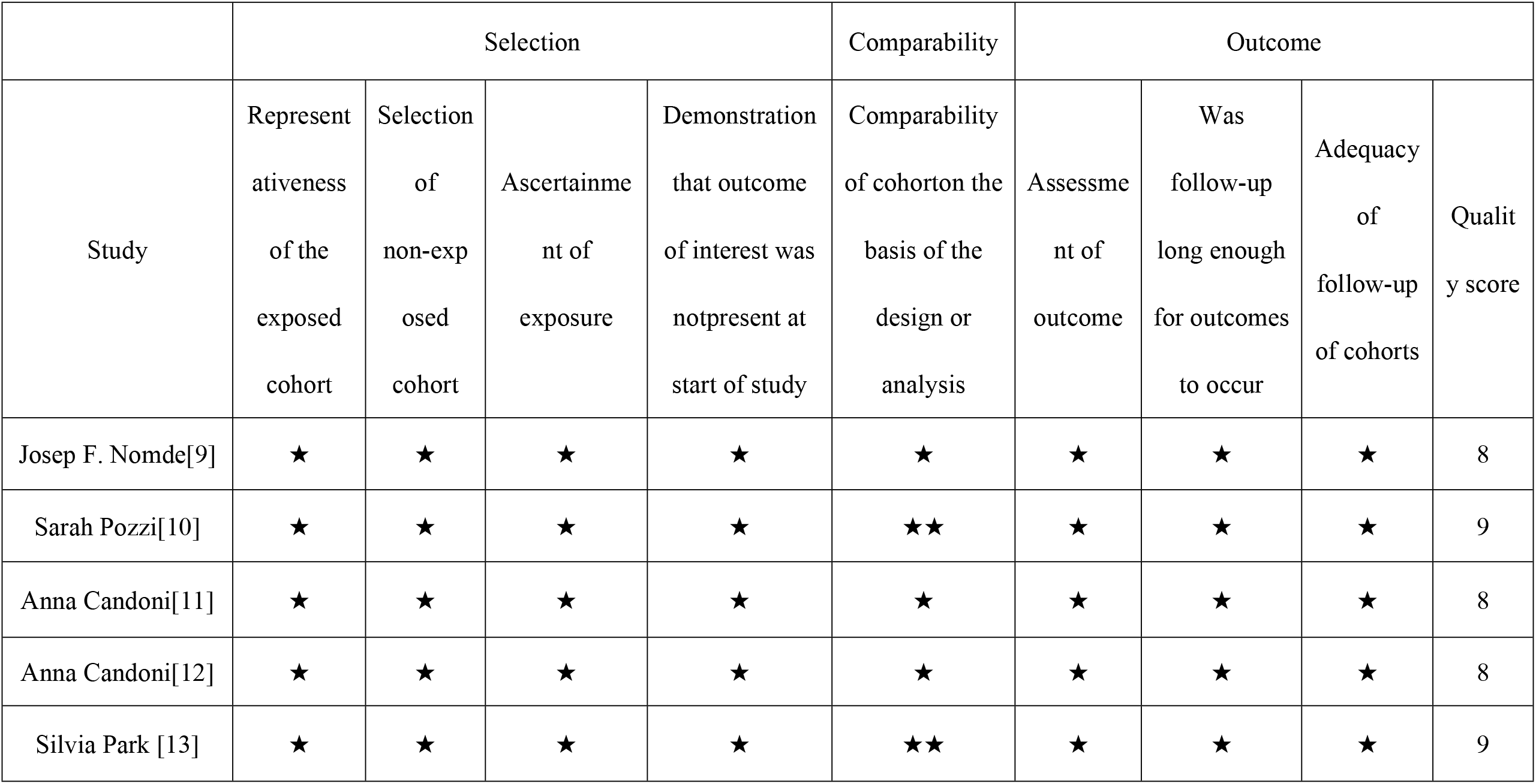
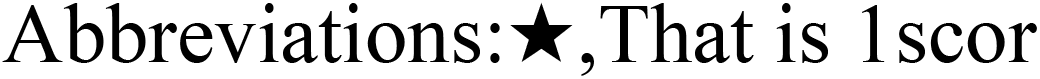
Results of quality assessment using the Newcastle-Ottawa Scale for case-control studies.

|  | Selection |  |  |  | Comparability | Outcome |  |  |  |
| --- | --- | --- | --- | --- | --- | --- | --- | --- | --- |
| Study | Representativeness of the exposed cohort | Selection of non-exposed cohort | Ascertainment of exposure | Demonstration that outcome of interest was not present at start of study | Comparability of cohort on the basis of the design or analysis | Assessment of outcome | Was follow-up long enough for outcomes to occur | Adequacy of follow-up of cohorts | Quality score |
| Josep F. Nomde[9] | ★ | ★ | ★ | ★ | ★ | ★ | ★ | ★ | 8 |
| Sarah Pozzi[10] | ★ | ★ | ★ | ★ | ★★ | ★ | ★ | ★ | 9 |
| Anna Candoni[11] | ★ | ★ | ★ | ★ | ★ | ★ | ★ | ★ | 8 |
| Anna Candoni[12] | ★ | ★ | ★ | ★ | ★ | ★ | ★ | ★ | 8 |
| Silvia Park [13] | ★ | ★ | ★ | ★ | ★★ | ★ | ★ | ★ | 9 |

Abbreviations:★,That is 1 scor

## Results of Meta-analysis

### WT1 expressed recurrence rate

RR was reported in four studies [10-13], and the random-effect model showed that compared with low expression, AML patients receiving Allo-HSCT with high expression had significantly higher 1-year RR [relative risk (RR) = 5.09, 95%CI (3.30, 7.83), *P* ≤ 0.001, I^2^ = 0.00%], and 2-year RR [RR = 4.09, 95%CI (2.71, 6.18), *P* ≤ 0.001, I^2^ = 24.83%], 4-year RR [RR = 3.27, 95%CI (2.42, 4.42), *P* ≤ 0.001, I^2^ = 0.00%], and 7-year RR (RR = 3.33, 95%CI (2.49, 4.46), *P* ≤ 0.001, I^2^ = 0.00%] (Figure 2A-D). It is suggested that AML patients after Allo-HSCT with WT1 high expression had a poor prognosis.

**Figure 2A:**
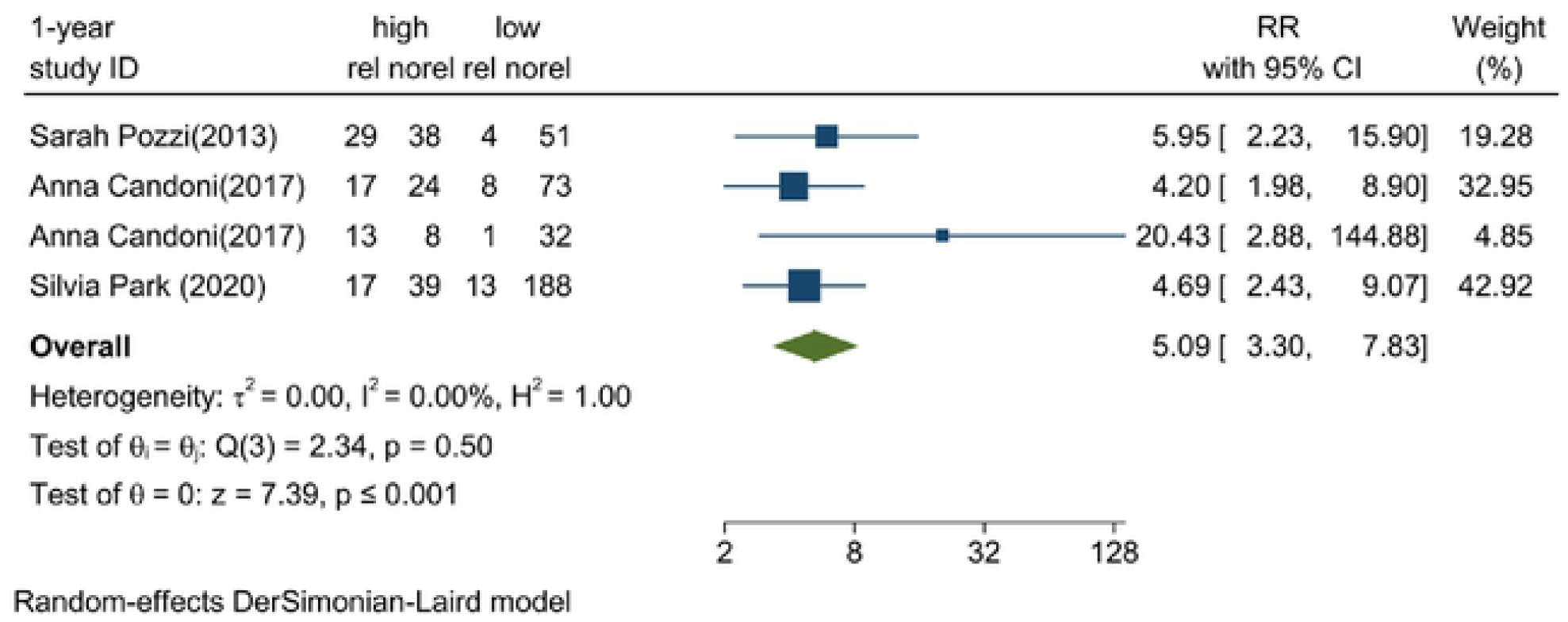
1-year recurrence rate.

**Figure 2B:**
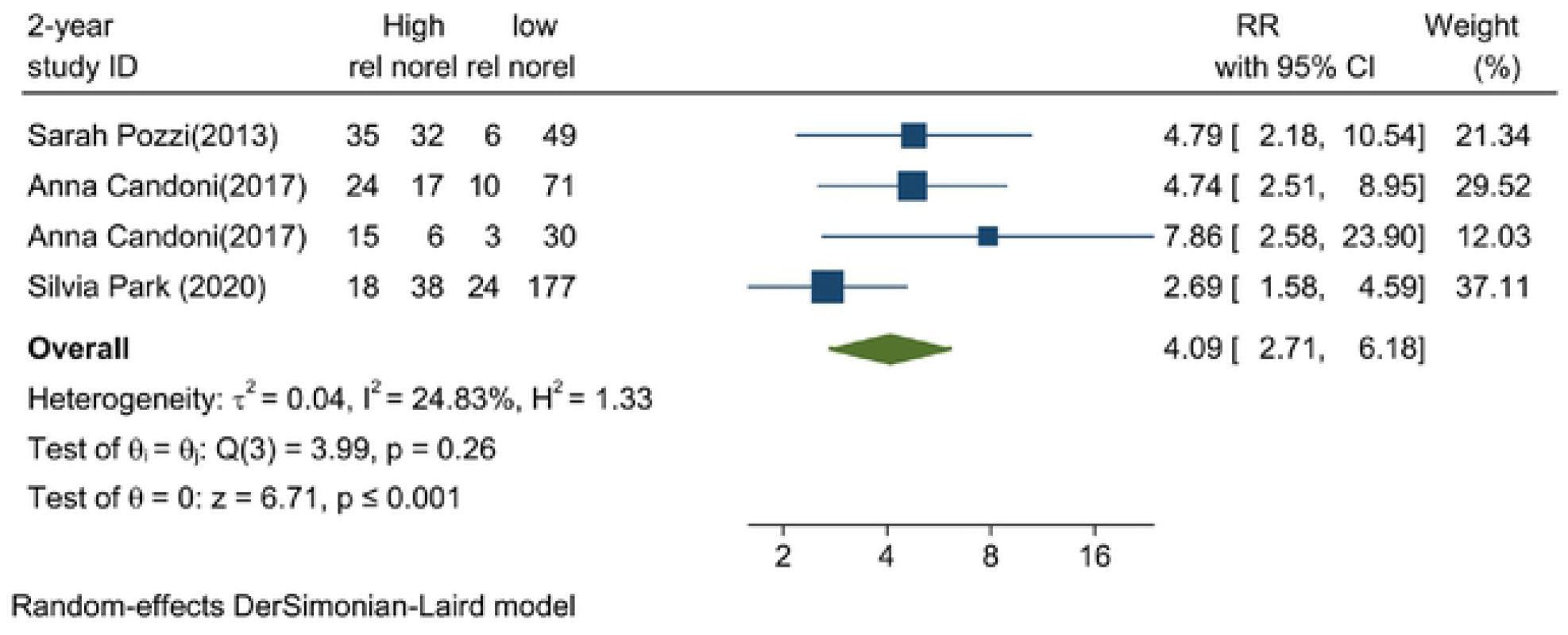
2-year recurrence rate.

**Figure 2C:**
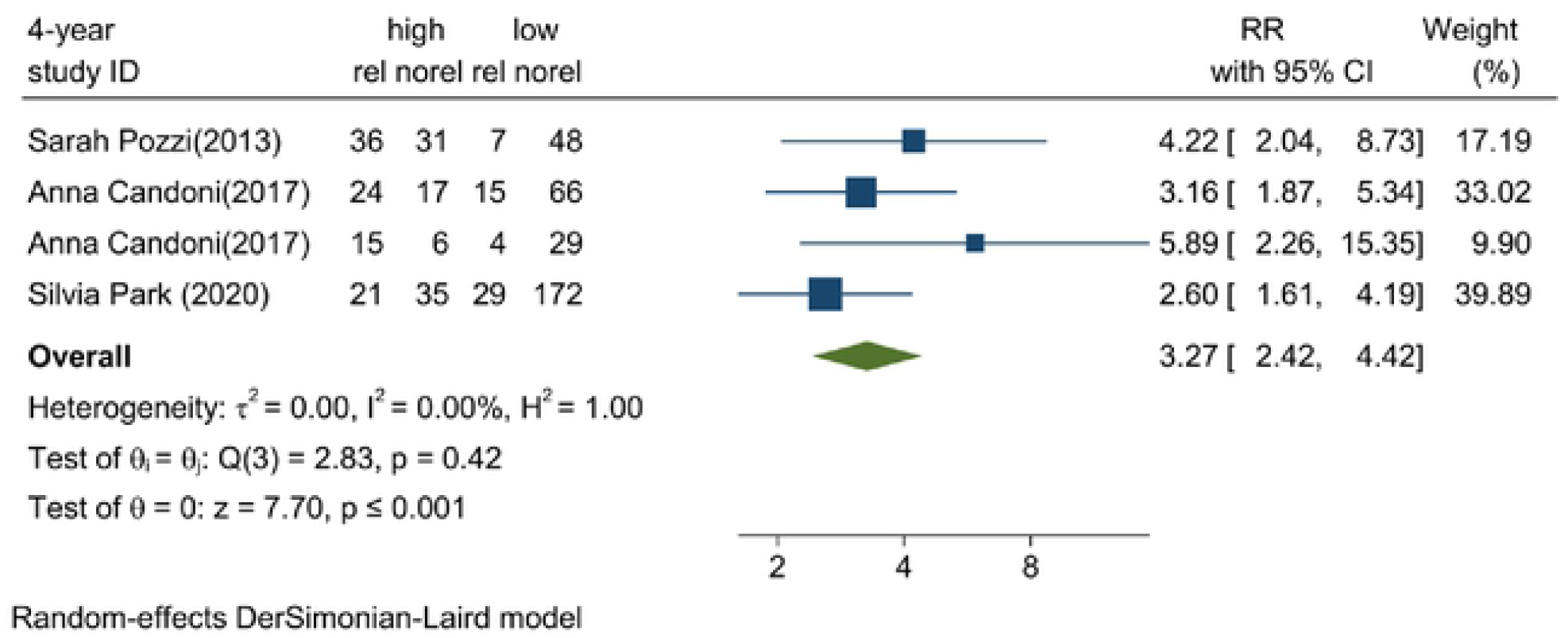
4-ycar recurrence rate.

**Figure 2D:**
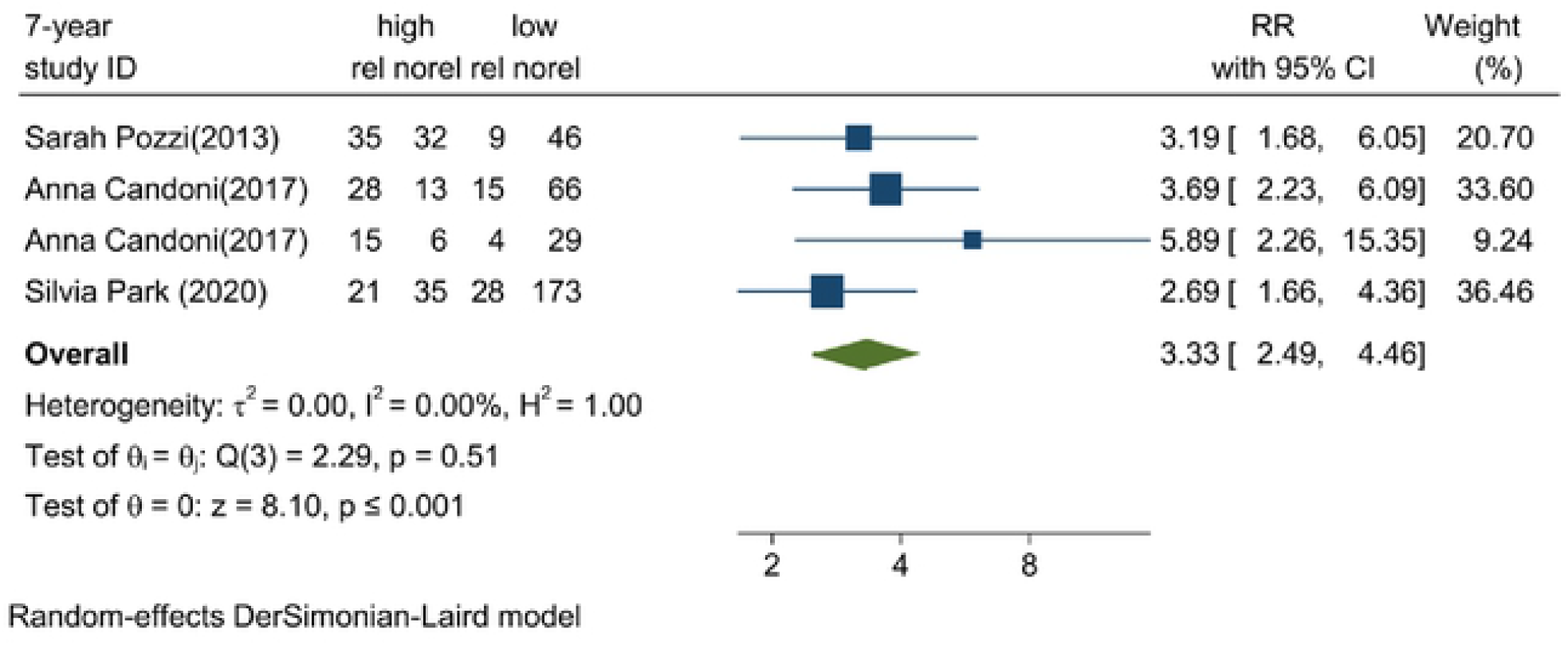
7-year recurrence rate.

### WT1 expression overall survival rate

OS was reported in four studies [10-13], and the random-effects model showed that the 1-year OS [RR=0.73, 95%CI (0.57, 0.94), *P*=0.02, I^2^=70.96%], 2-year OS [RR=0.59, 95%CI (0.45, 0.77), *P* ≤ 0.001, I^2^=53.53%], 4-year OS [RR = 0.54, 95%CI (0.38, 0.75), *P* ≤ 0.001, I^2^ = 55.88%] and 7-year OS [RR = 0.51, 95%CI (0.37, 0.70), *P* ≤ 0.001, I^2^ = 35.13%] were significantly higher low expression than high expression (Figure 3A-D). It is suggested that AML patients after Allo-HSCT with low expression had a better prognosis.

**Figure 3A:**
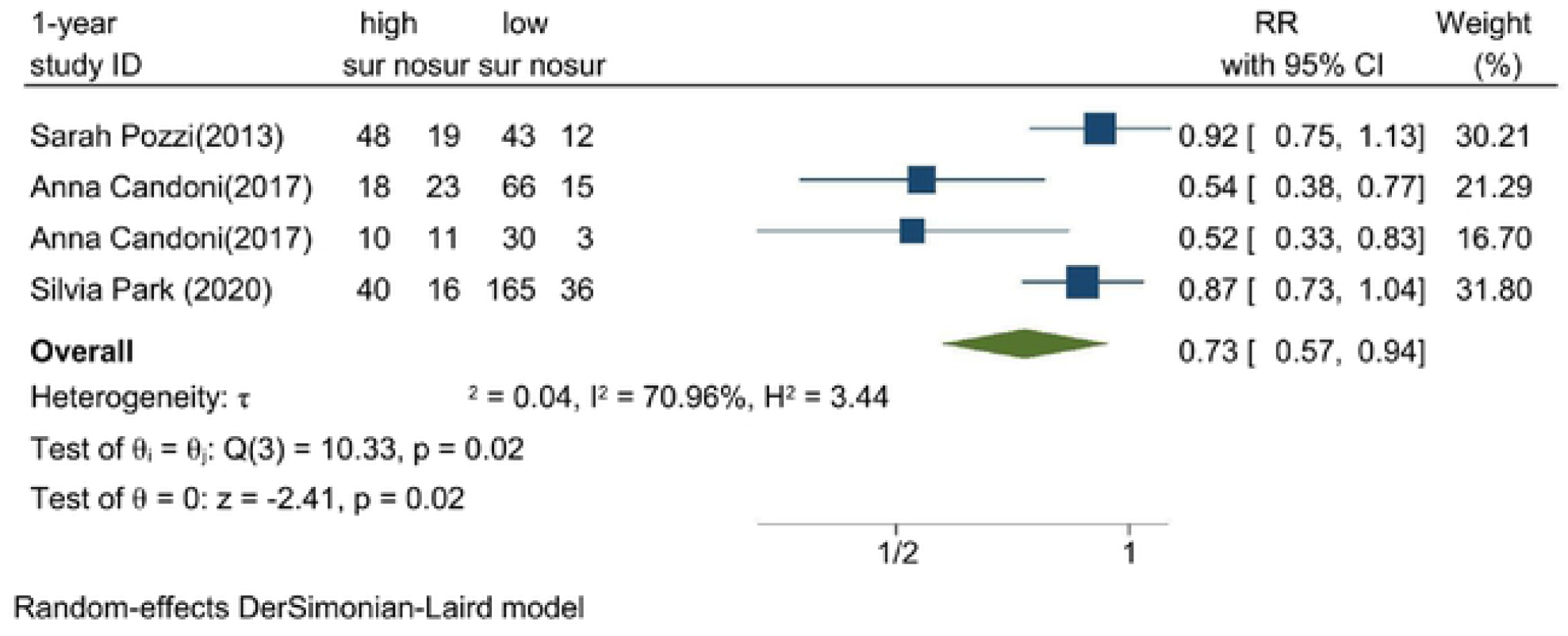
1-year overall survival.

**Figure 3B:**
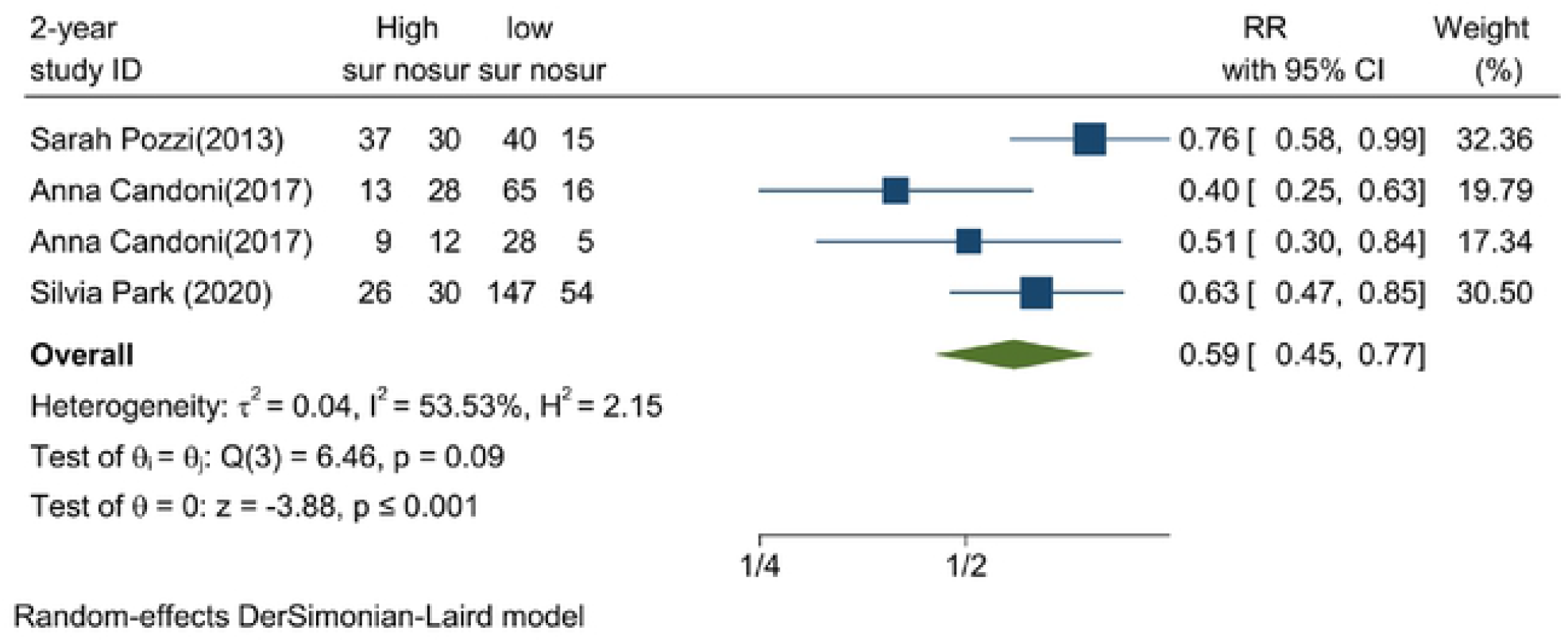
2-year overall survival.

**Figure 3C:**
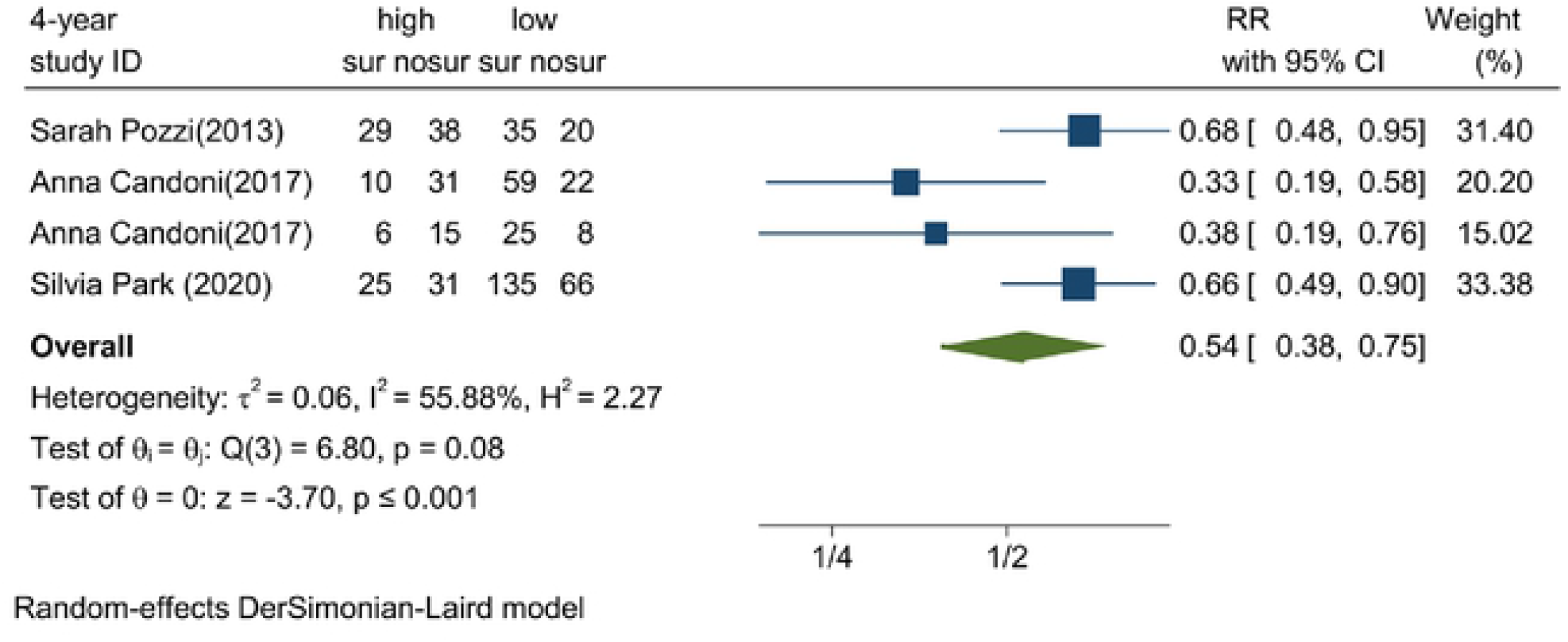
4-year overall survival.

**Figure 3D:**
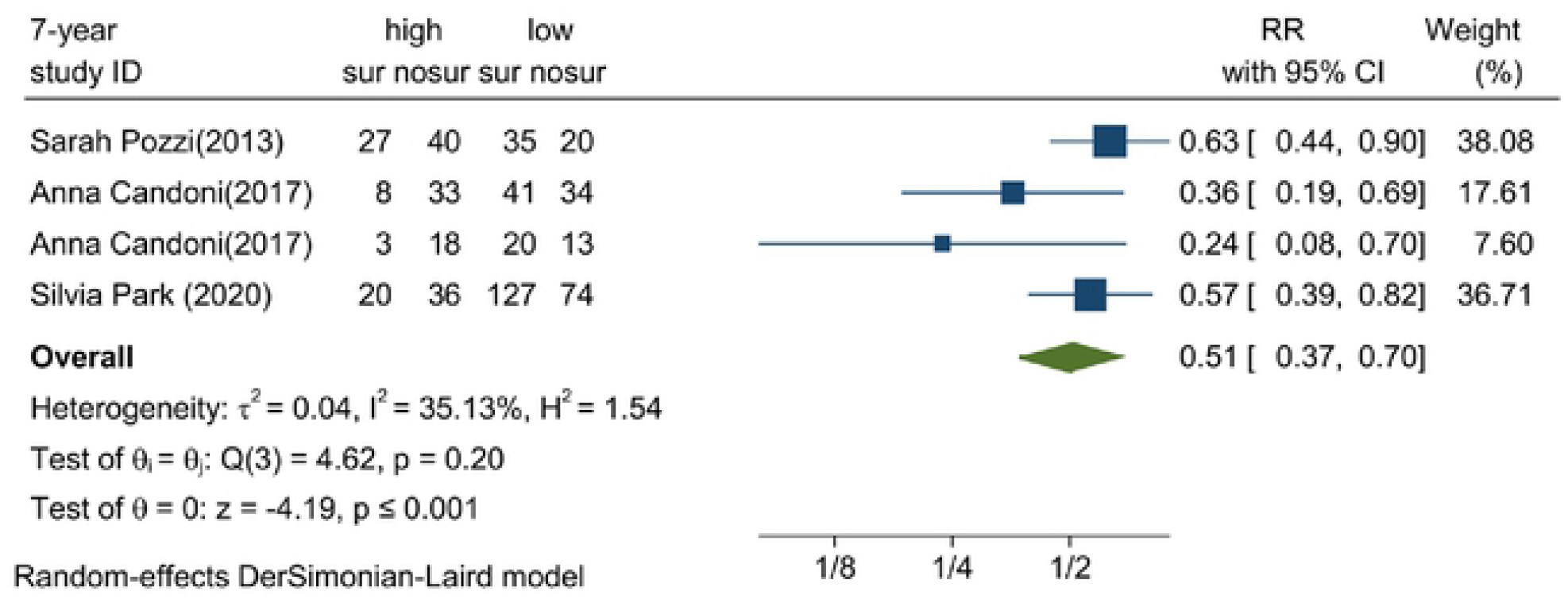
7-year overall survival.

### The disease-free survival rate of experimental group and control group

Three studies reported DFS [9,11,12], and the random-effects model showed that the 1-year DFS was significantly lower in experimental group than that of control group [RR = 1.19, 95%CI (1.03, 1.38), *P* = 0.02], while no significant difference was detected in the 4-year DFS [RR = 1.18, 95%CI (0.98, 1.42), *P* = 0.09] (Figure 4A, B).

**Figure 4A:**
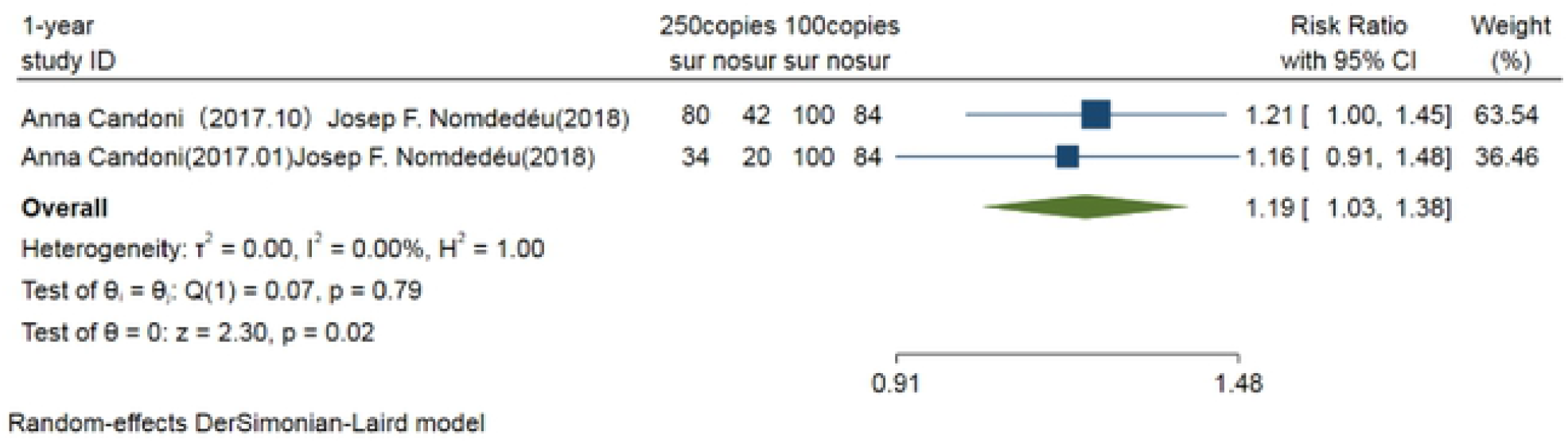
1-year disease-free survival.

**Figure 4B:**
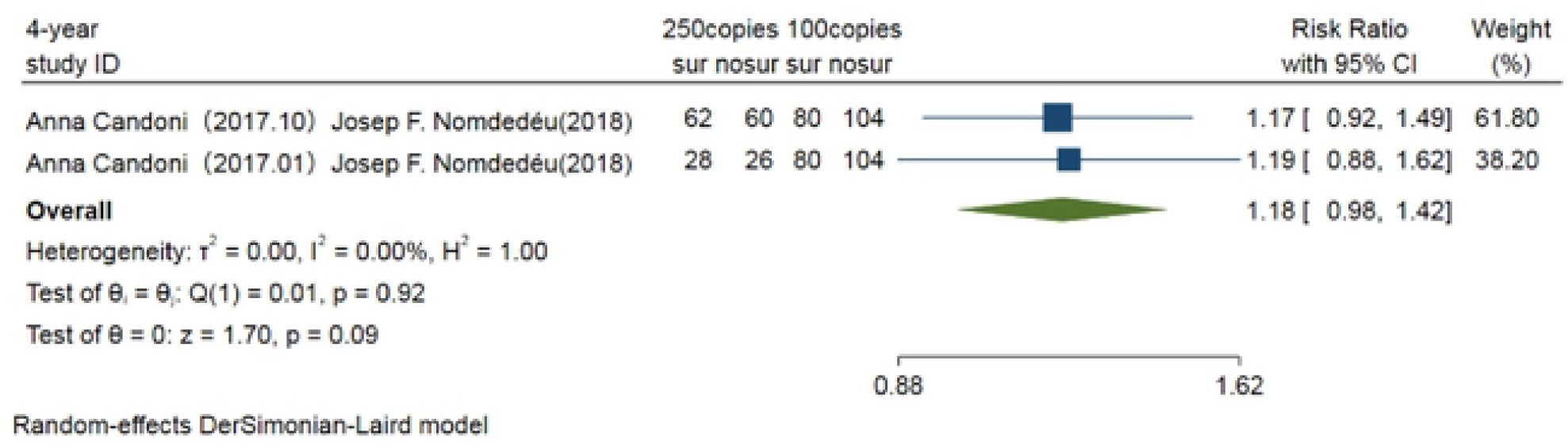
4-year disease-free survival.

### The overall survival of experimental group and control group

OS was reported in four studies [9-12], and compared in experimental group and control group. The random-effects model showed that the 4-year OS in experimental group was significantly lower than that of control group [RR = 1.16, 95%CI (1.03, 1.32), *P* = 0.02], while no significant difference was detected in the 1-year OS between experimental group and control group [RR = 1.06, 95%CI (0.92, 1.23), *P* = 0.40] (Figure5A, B). It is suggested that the threshold of WT1 at 250 copies was an appropriate threshold for predicting the poor prognosis of AML after Allo-HSCT.

**Figure 5A:**
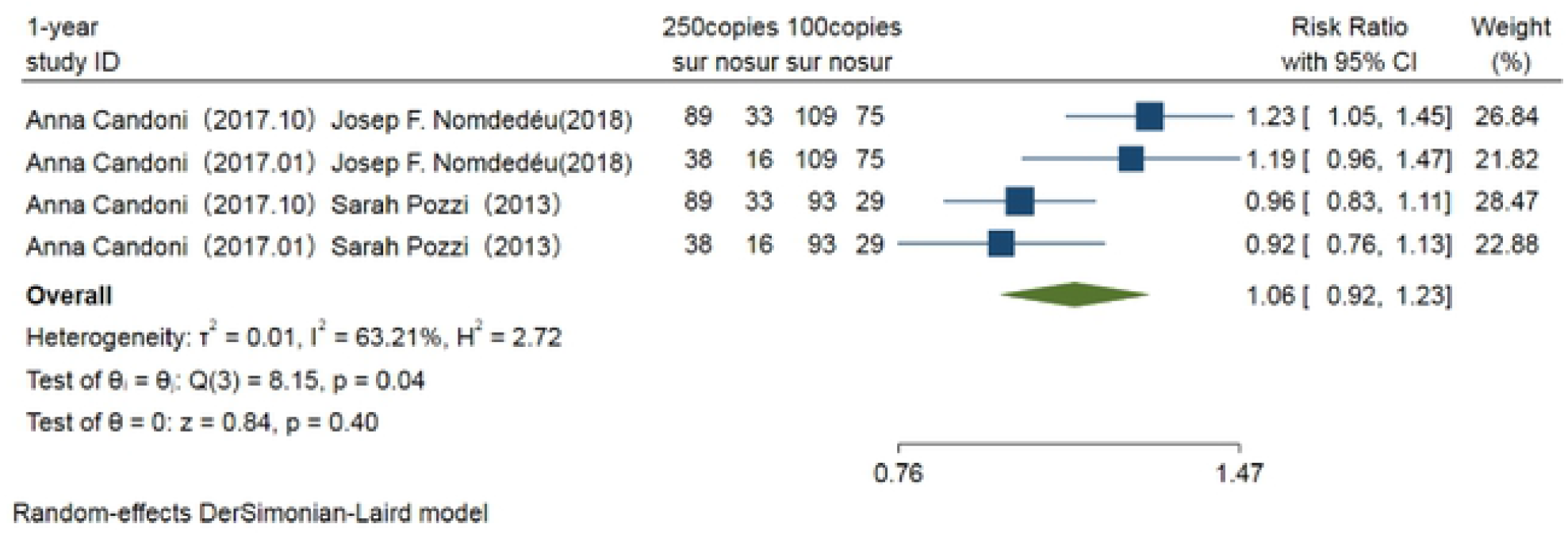
1-year overall survival.

**Figure 5B:**
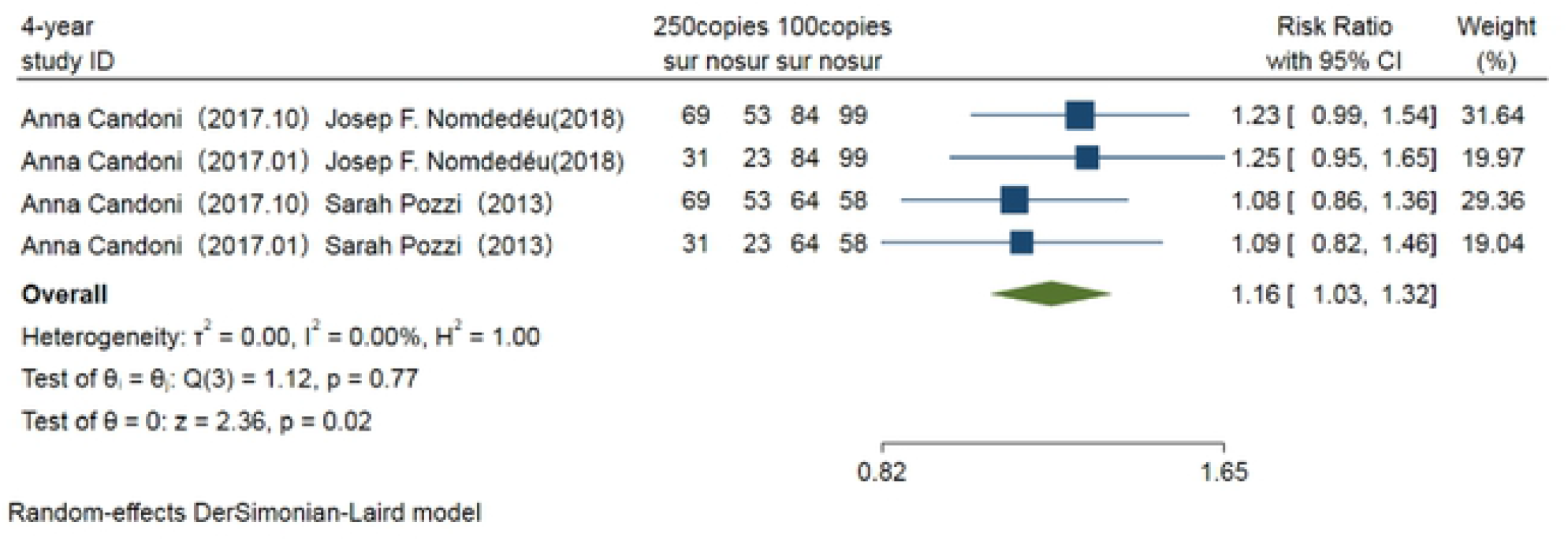
4-year overall survival.

### Heterogeneity analysis

The 1-year, 2-year, 4-year, and 7-year OS analysis of high expression and low expression was performed by introducing the random-effects model. However, I^2^≥50% suggested heterogeneity. We subsequently explored the possible causes of heterogeneity through subgroup analyses based on the country and threshold of WT1. Subgroup analysis on 2-year OS showed that I^2^ = 32.95%, suggesting that the 2-year OS may be the cause of heterogeneity. In addition, we conducted sensitivity analysis by excluding one literature each time. It is shown that I^2^ = 14.92% after excluding the literature reported by Candoni et al. [11] that reported 4-year OS, and I^2^ = 12.44% after excluding the literature reported by Candoni et al. [12] that reported 7-year OS, indicating that data extracted from literature of Candoni et al. [11] and Candoni et al. [12] were sources of heterogeneity. The heterogeneity of 1-year OS may be related to publication bias. Although there was significant heterogeneity in 1-year, 2-year, 4-year and 7-year OS between high expression and low expression, the above factors affected the reliability of OS through heterogeneity analysis.

### Publication bias

Stata 16.0 software was used for depicting funnel plots and the Egger test was performed to evaluate publication bias. RR of AML patients after HSCT in high expression and low expression were analyzed, and funnel plots were well symmetric. Egger test results showed that there were no significant differences in the 1-year (*P* = 0.143), 2-year (*P* = 0.078), 4-year (*P* = 0.099) and 7-year RR (*P* = 0.238) of AML patients between the high expression and low expression, suggesting no publication bias. In addition, there were significant differences in the 1-year (*P* = 0.002), 2-year (*P* = 0.025), 4-year (*P* = 0.016) and 7-year OR (*P* = 0.036), suggesting the publication bias in the included studies. It is considered that authors, research sponsors and manuscript editors are confounding factors for data reliability. The funnel plots of disease-free survival rate and overall survival rate in the 250 copies /104ABL experimental group and the 100 copies /104ABL control group showed good symmetry, as shown in Fig. 6A and B, and Fig. 7A and B. There were no significant differences in the 1-year DFS (*P* = 0.812), 4-year DFS (*P* = 0.938), 1-year OS (*P* = 0.875), and 4-year OS (*P* = 0.968), suggesting no publication bias.

**Figure 6:**
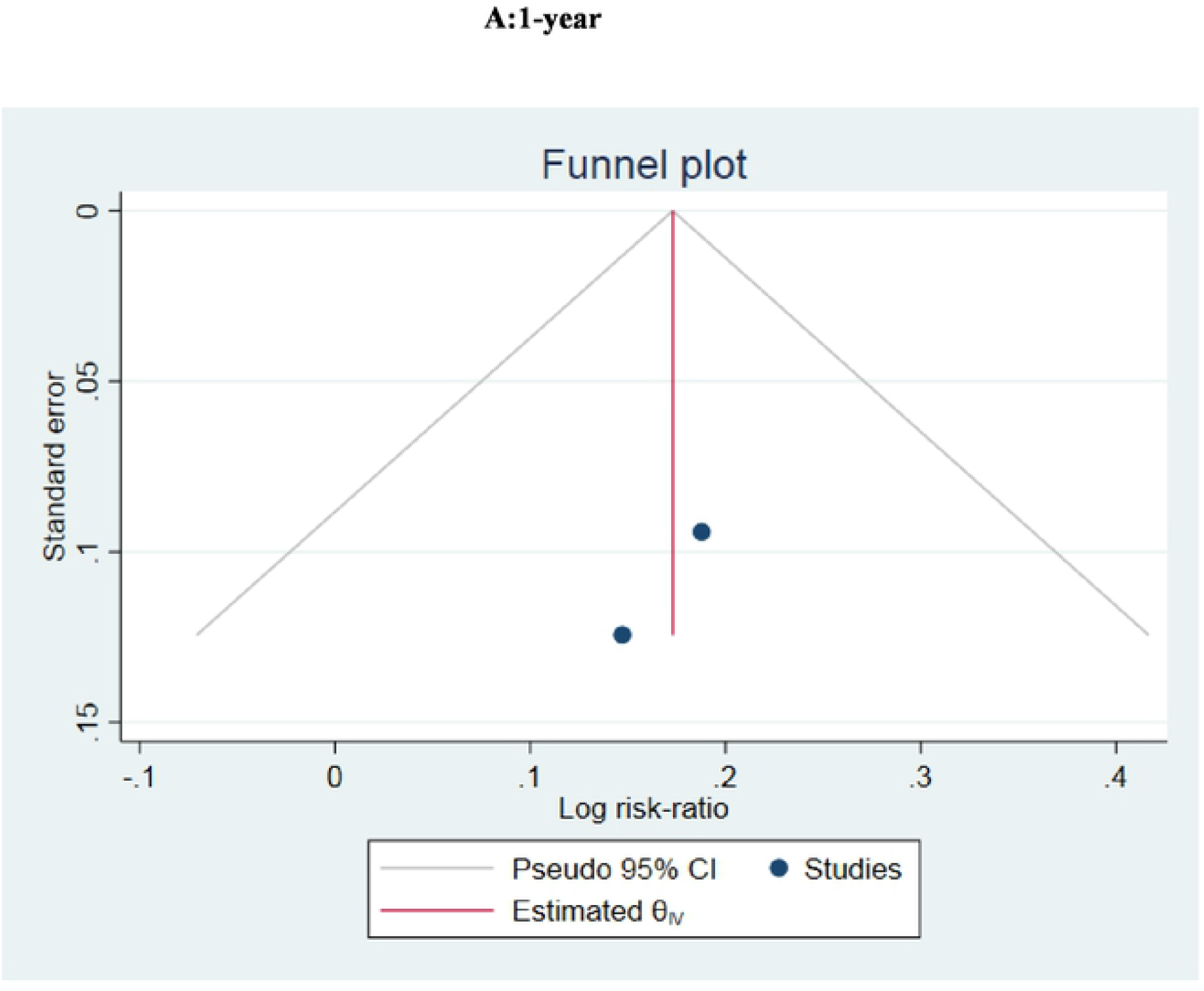
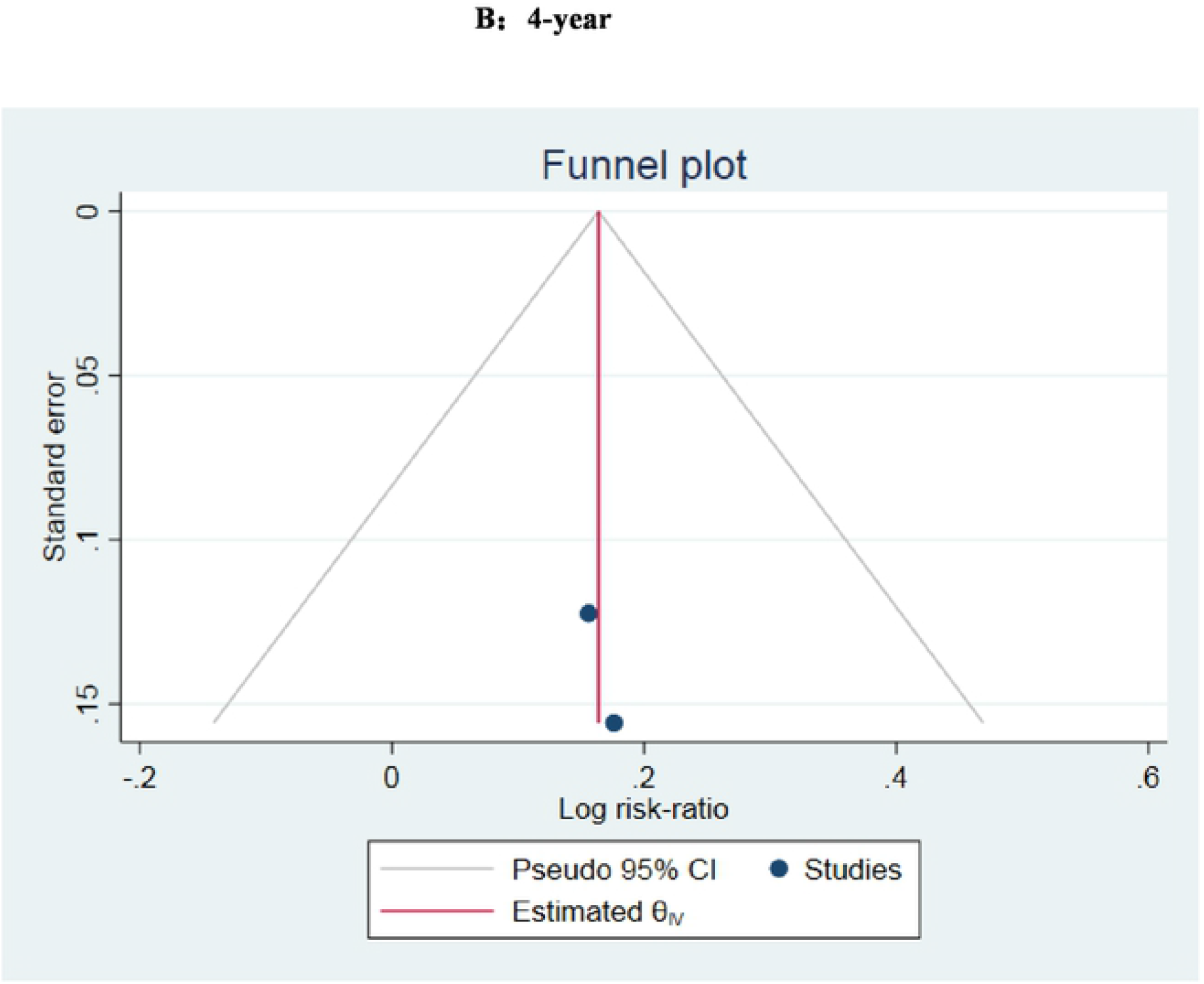
Funnel plot of disease free survival rate.

**Figure 7:**
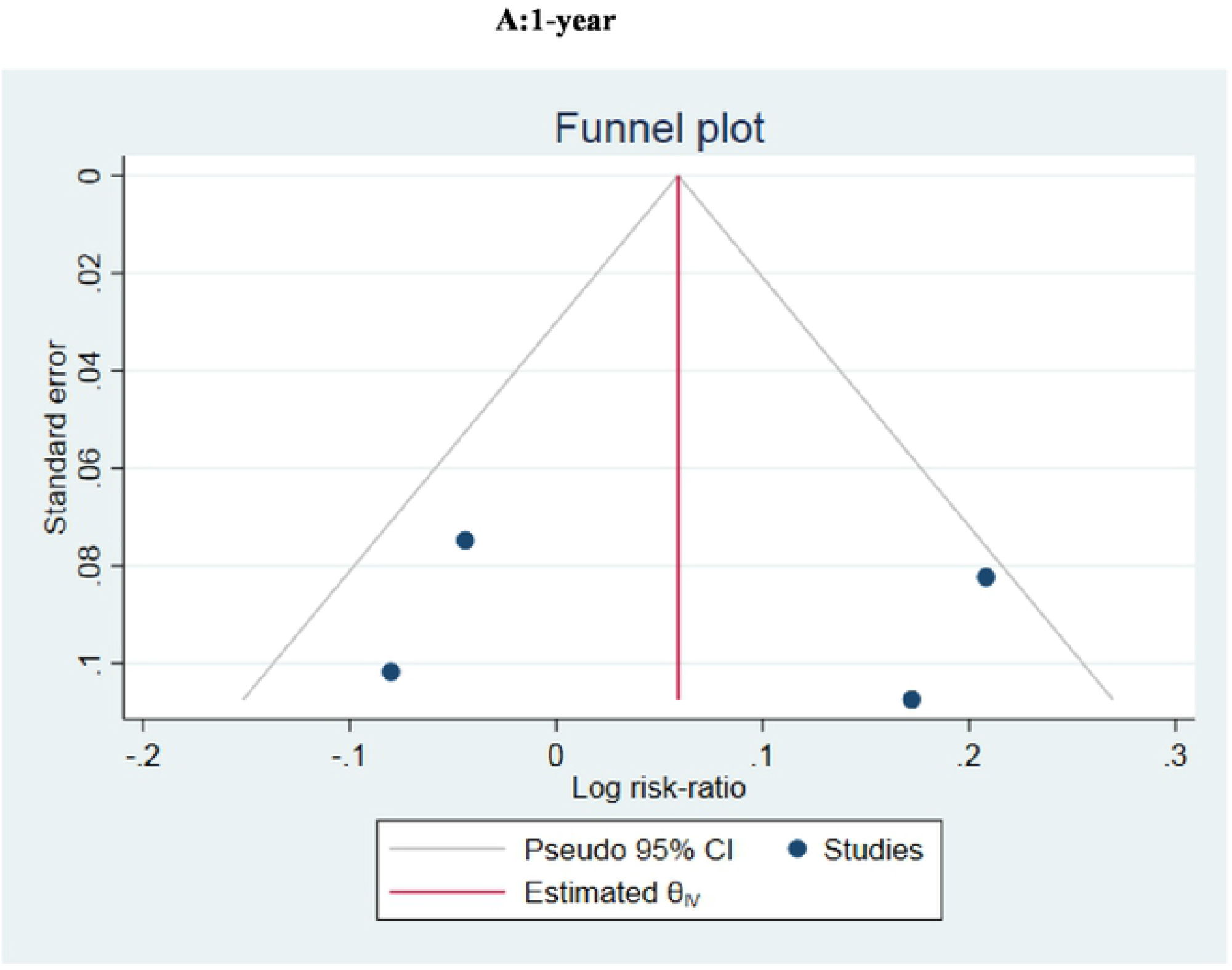
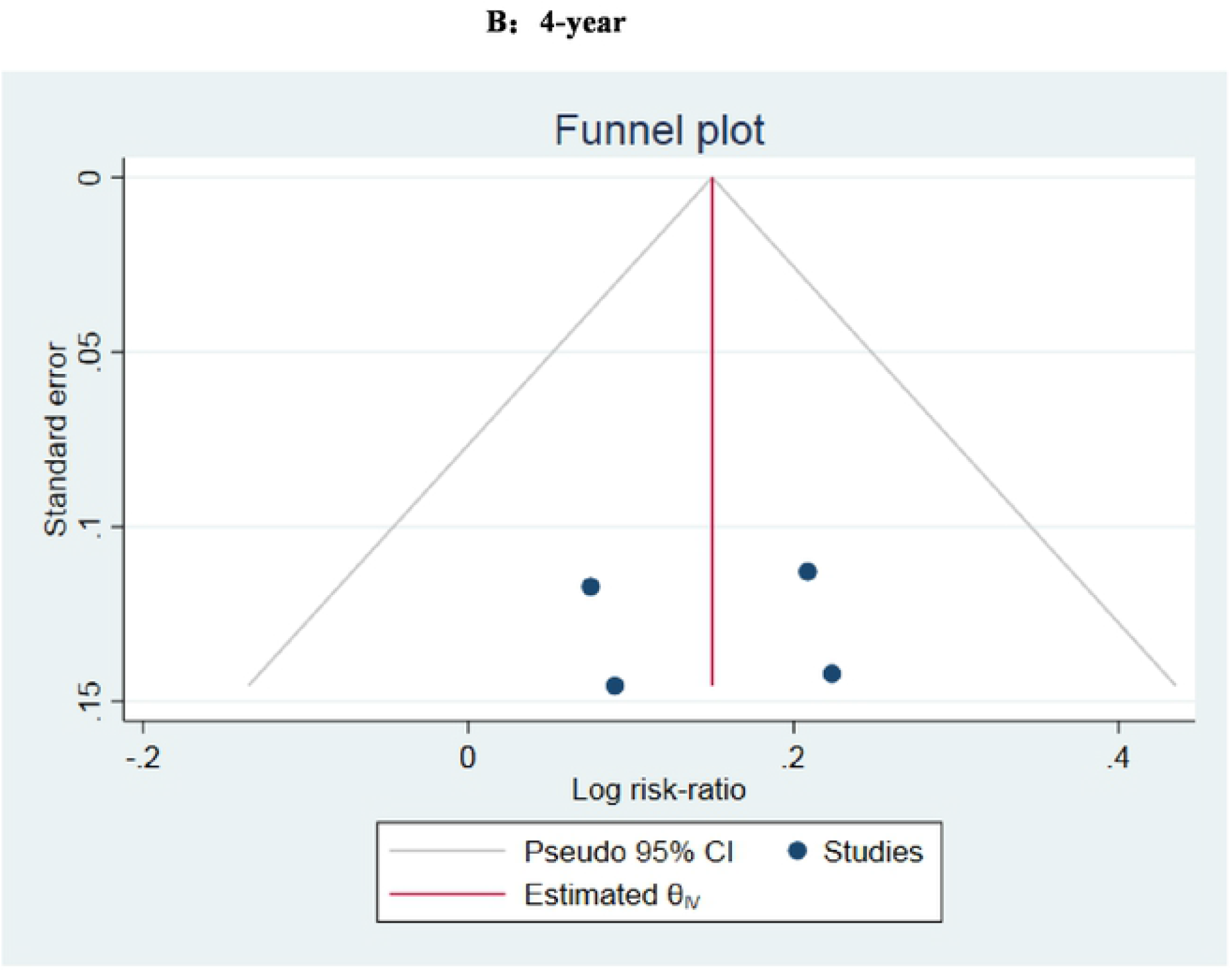
Funnel plot of overall survival.

## Discussion

The WT1 gene is located on chromosome 11PL3, which was originally identified as a tumor-suppressor gene associated with the Wilms tumor, a childhood kidney cancer. WT1 is overexpressed in various solid cancers and hematological malignant tumors [14]. Moreover, it is overexpressed in bone marrow (BM) or peripheral blood (PB) of more than 80% of AML patients, and is also served as a strong and independent marker of minimal residual disease (MRD) [15]. Multiple measurement methods of WT1 expression level in PB and BM have been proposed as follows [16,17, 19-24]. (1) Ratio method: Real-time quantitative PCR (RT-qPCR), with ABL gene as the internal reference in the unit of 104 ABL copies; (2) Logarithmic method: Logarithmic order of WT1 gene decline; (3) Standard curve method: The mRNA level of WT1 detected in 1 μg K562 cells was defined as 1 to normalize its level in samples. In our included papers, ratio method was used to measure the expression level of WT1 in BM. Besides, real-time quantitative polymerase chain reaction (RT-qPCR) was used to measure mRNA levels of WT1 and ABL according to the European Leukemia Network (ELN), and the mRNA level of WT1 was normalized to 104 ABL copies as the unit. Here, ratio method was used to evaluate the effect of WT1 expression level on the prognosis of AML patients receiving Allo-HSCT. Recently, quantitative analysis of the WT1 gene combined with multi-parameter flow cytometry has significantly improved the sensitivity in predicting the risk of AML recurrence after Allo-HSCT, which has been applied in analyzing the prognosis of AML patients after Allo-HSCT [18].

In 2009, Cilloni et al. [19] defined overexpressed WT1 based on ELN as ≥ 50 copies /104 ABL copies in peripheral blood samples and ≥ 250 copies / 104 ABL copies in bone marrow samples detected by RT-qPCR. In other studies, ratio method is also used to evaluate the effect of WT1 expression on disease prognosis. Cho et al. [20] tested WT1 in BM samples in patients after Allo-HSCT, and suggested that 250 copies of WT1 is the optimal threshold and 3 months after Allo-HSCT was the optimal time point to detect the copy number of WT1. Frairia et al. [21] believed that WT1 > 150 copies / 104 ABL copies in BM samples before transplantation is independently associated with RR and survival. In this study, the WT1 threshold was different from the ELN threshold. Sun et al. [22] believed that WT1 > 87 copies /104 ABL copies in BM samples indicates a poor prognosis. Zhao et al. [23] believed that WT1 > 585 copies /104 ABL copies in BM samples is unfavorable to the prognosis after transplantation. In addition, Mashima et al. [24] found that two consecutive tests of WT1 transcription levels at more than 100 copies during remission may help identify patients at high risk of relapse. threshold of WT1 expression are inconsistent in many studies, which limits its routine clinical application. Two reasons may explain the inconsistent findings. Firstly, AML is a highly heterogeneous disease composed of subgroups with distinct biological characteristics [25]. Secondly, ELN threshold or threshold calculated based on the median of WTl threshold is adopted in different studies [8]. Due to the lack of relevant guidelines and consensus on the optimal threshold of WT1 expression, this study analyzed the prognostic factors for AML patients after Allo-HSCT by meta-analysis. It is shown that 1-year DFS in the experimental group was significantly lower than that in the control group. The 4-year OS of the experimental group was significantly lower than that of the control group, suggesting that the threshold of 250 copies/104 ABL of WT1 may be the optimal threshold for predicting the poor prognosis in AML patients after Allo-HSCT. In the heterogeneity analysis, data heterogeneity was found in Candoni et al. [11] and Candoni et al. [12], and 250 copies /104 ABL was defined as overexpression in these two papers. Therefore, considering the accuracy of the results of this study, TSAV0.9 software was used for sequential analysis to evaluate the sample size of meta-analysis. The results are shown in Figure 8A and B, which showed that this study did not reach the expected sample size (TSAn=5064) and there were false positive results. Therefore, 250 copies /104 ABL of WT1 expression as the optimal threshold should be further expanded to provide strong evidence for the impact on prognosis.

**Figure 8:**
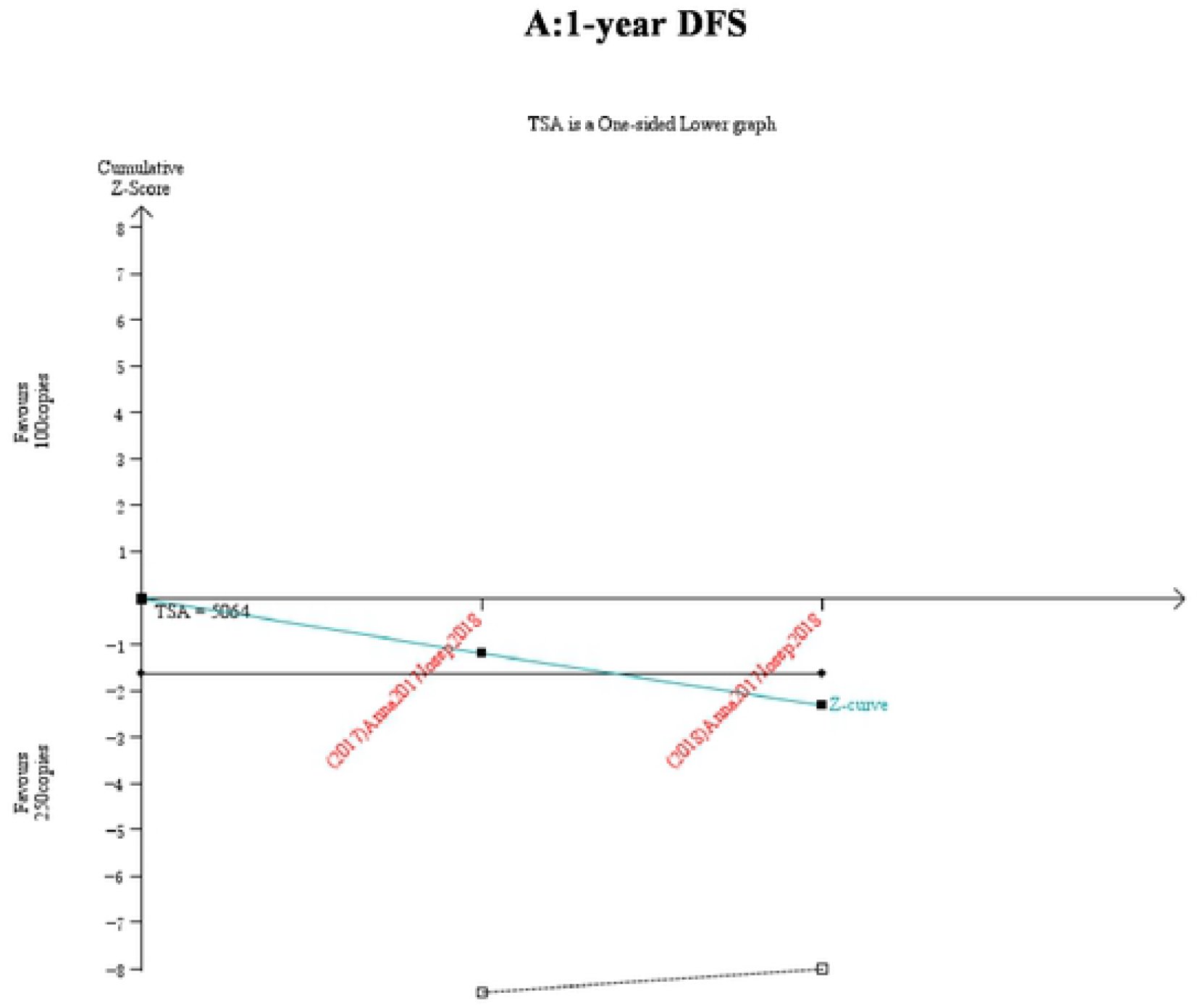
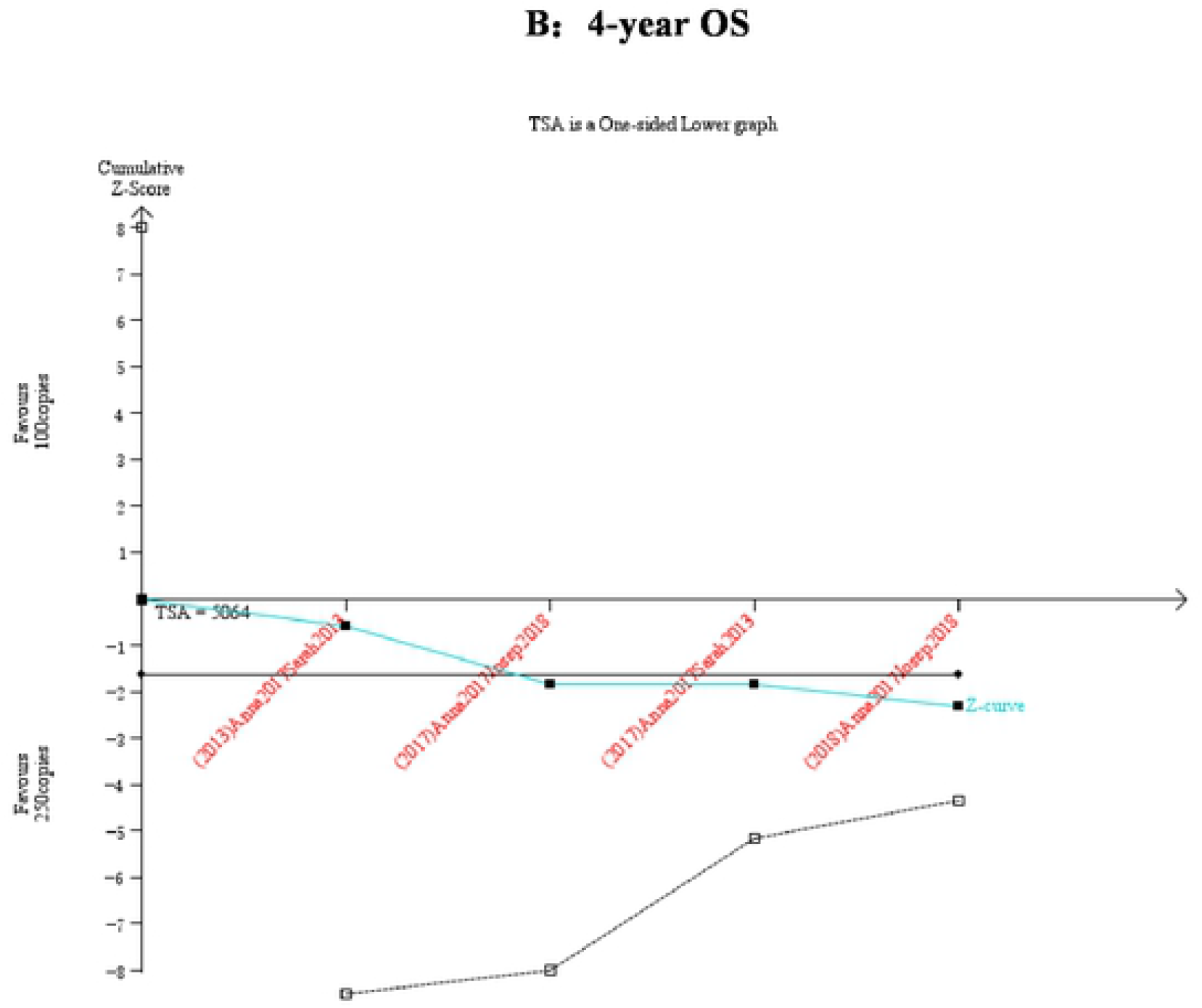
The TSA curve.

Previous studies have not highlighted the comparison between different thresholds of WT1 expression, and its optimal threshold. This study compared the effects of two different WT1 expression thresholds on the prognosis of AML patients after Allo-HSCT, so as to provide evidence for future clinical application. WT1 is overexpressed in a variety of hematopoietic tumors like AML, and previous studies have found that upregulated WT1 is associated with leukemia recurrence [16]. AML patients after Allo-HSCT expressing a high level of WT1 generally have a higher risk of recurrence, which negatively affects the survival. In this study, AML patients with high WT1 expression after Allo-HSCT were followed up at 1, 2, 4 and 7 years. Compared with patients with low WT1 expression of AML after Allo-HSCT, patients with high WT1 expression had a higher recurrence rate and a higher overall survival rate, which was similar to the results of previous published studies.

The optimal treatment for AML has not yet been established, and Allo-HSCT is the first-line treatment for low-risk and very low-risk AML patients [26], while post-transplant recurrence is a poor prognostic factor. Multiple studies have shown that monitoring WT1 expression level after Allo-HSCT can predict its recurrence [23]. In the present study, the cut-off value at 250 copies of WT1 was found to be the optimal threshold for predicting the poor prognosis of AML after Allo-HSCT. Monitoring the optimal overexpression threshold of WT1 contributes to the early identification of the recurrence and timely performance of donor lymphocyte infusion in AML patients after Allo-HSCT [25]. In addition, immunotherapy targeting WT1 may have the potential to prevent recurrence and improve survival [27]. Chapuis et al. [28] found that T cell receptor gene therapy targeting WT1 can prevent the recurrence of AML after transplantation. Recent studies have shown that WT1 peptide vaccine can inhibit tumor growth, and postoperative and/or chemotherapy vaccination can significantly reduce disease recurrence in patients with high tumor load [29]. WT1 gene expression may be related to the prognosis of AML with other factors. For example, the transplantation does not achieve disease remission or the presence of high-risk chromosomal abnormalities. Hidaka et al. [30] reported that WT1 expression is higher in INV (16) and T (15:17)AML patients than in T (8:21)AML patients, and FLT3-ITD or NPM1 mutation is independently associated with high WT1 expression in AML patients. A variety of WT1 gene mutations have been reported in solid tumors and AML. Wang et al. [31] believed that WT1 mutation is an independent adverse prognostic factor in pediatric AML.

Several limitations in this study should be noted. (1) Due to the small sample size, this may reduce the statistical power of the results, and further expansion of the sample size is needed. (2) The lack of patients’ data and the limited number of studies may result in potential heterogeneity that influenced the accuracy of the results. (3) Chinese and English published literatures were searched in this study, but the classification of the threshold was only reported in foreign countries, which may have a certain publication bias.

In conclusion, high expression of WT1 significantly affects the prognosis of AML patients after Allo-HSCT, which is served as an adverse factor for predicting the recurrence. In detail, the threshold at 250 copies/104ABL of WT1 is the optimal threshold for predicting the adverse prognosis of AML after Allo-HSCT. It is recommended to closely monitor the expression level of WT1 based on the calculated optimal threshold of copy numbers, thus predicting the recurrence, reducing adverse events and improving clinical outcomes in AML patients after Allo-HSCT. Due to the small sample size, it is necessary to further expand the sample size to validate our findings.

## Data Availability

All relevant data are within the manuscript and its Supporting Information files.

No

## Acknowledgments

Thanks for the support of translation Center of Henan University of Science and Technology.

## Conflict of interests

The authors declare that there is no conflict of interests regarding the publication of this paper.

## Availability of data

All data were extracted from the original articles included.

## Notes

### Competing Interest Statement

The authors have declared that no competing interests exist.

### Clinical Trial

No

### Clinical Protocols

No

### Funding Statement

The author received no specific funding for this work.

